# QMRA of SARS-CoV-2 for workers in wastewater treatment plants

**DOI:** 10.1101/2020.05.28.20116277

**Authors:** Rafael Newton Zaneti, Viviane Girardi, Fernando Rosado Spilki, Kristina Mena, Ana Paula Campos Westphalen, Evandro Ricardo da Costa Colares, Allan Guedes Pozzebon, Ramiro Gonçalves Etchepare

## Abstract

Faecal-oral transmission of SARS-CoV-2 is a hot topic and additional research is needed to elucidate the risks of the novel coronavirus in sanitation systems. This is the first article that investigates the potential health risks of SARS-CoV-2 in sewage to wastewater treatment plants (WWTPs) workers. A quantitative microbial risk assessment (QMRA) is applied for three COVID-19 scenarios (moderate, aggressive and extreme) to study the effect of different stages of the pandemic, in terms of percentage of infected population, on the probability of infection. Results reveal that estimates of viral loads in sewage at the entrance of WWTPs ranged from 1.03×10^2^ to 1.31×10^4^ GC.mL^−1^ (0.1 to 13.06 PFU.mL^−1^, respectively) and that estimated risks for the aggressive and extreme scenarios (6.5×10^−3^ and 3.1×10^−2^, respectively) were likely to be above a WHO benchmark of tolerable risk used for virus infection of 10^−3^ and higher than the risk of infection of *E. coli*, used herein as common pathogen indicator for a relative comparison, thus reinforcing the concern of sewage systems as a transmission pathway of SARS-CoV-2. These findings are helpful as an early-warning tool and in prioritizing upcoming risk management strategies in the sanitation sector during COVID-19 pandemic.

## 1. Introduction

The primary mechanism of SARS-CoV-2 transmission is via respiratory droplets that people cough, sneeze or exhale (ECDPR, 2020). The virus can survive on different surfaces from several hours up to a few days, and although its persistence in waters is possible, the stability of its infectivity is not fully understood. Human viruses do not replicate in the environment and the transport of SARS-CoV-2 via the water cycle fosters its ability to survive in human wastewater catchments, by retaining its infectivity and coming in contact with people, most likely via aerosols and airborne particles (Haas, 2020; McLellan at al., 2020; Li et al., 2020). Diarrhea, as reported in a great proportion of the COVID-19 cases, appears to have a direct influence on a secondary path of transmission for SARS-CoV-2 as many recent peer-reviewed articles and reports have shown that the virus has been detected in stool samples of COVID-19 patients (Mao et al., 2020; Kitajima et al., 2020; Wu et al., 2020; Holshue et al., 2020, Murphy and Whitfield, 2020; Pan et al., 2020; Lescure et al., 2020; Zhang et al., 2020; Wang et al., 2020). Pan et al. (2020) examined stool samples from 17 COVID-19 cases by an N-gene-specific quantitative (RT-qPCR) assay and reported that 53% of the individuals were positive using RT-qPCR analysis. Corroborating, Lescure et al. (2020) followed the patterns of clinical disease and viral load of five patients diagnosed with COVID-19 by RT-qPCR analysis. Their results showed that two out of five patients had a positive detection of SARS-CoV-2 in stool samples with an average viral load of 6.61×10^7^ genomic copies (GC).g^−1^ of stool. Yet, SARS-CoV-2 shedding was investigated in a group of nine COVID-19 cases over a three-week period. In this study, viral RNA was detected in faeces, ranging in concentrations from 10^7^ RNA copies.g^−1^, in the first week, to 10^2^ RNA copies.g^−1^, in the third week of symptoms (Wölfel et al., 2020). Accordingly, the study of Zhang et al. (2020) analyzed the faeces of 19 COVID patients and the median duration of virus shedding was 22 days. Importantly, the first confirmation of infectious SARS-CoV-2 in faeces has been reported by Xiao et al (2020).

In light of the ability of SARS-CoV-2 to remain viable in conditions that would facilitate transmission via fecal-oral, it is possible that SARS-CoV-2 can also be transmitted through this route under special conditions, as suggested by a few authors quite recently (Yeo et al., 2020; Heller et al., 2020; Kitajima et al., 2020). Thus, the possibility of fecal-oral transmission of SARS-CoV-2 has many implications, especially in regions with poor sanitation infrastructure, considering the possible entering of SARS-CoV-2 into the sewage and wastewater treatment plants (WWTPs). After the first SARS global outbreak of 2003, very little information was available on the presence of SARS-CoV in sewage, with the exception to the study by Wang et al. (2005), which reported that SARS-CoV could be excreted through the stool/urine of infected patients into the sewage system and remain infectious for 2 days at 20 °C, but for 14 days at 4 °C, thus demonstrating the sewage system as possible route of transmission. In the current COVID-19 pandemic, Medema et al. (2020) published the first report of detection of SARS-CoV-2 in WWTPs, by analyzing (using RT-PCR) sewage samples from seven different cities and from an airport in the Netherlands, during a period before and after the first COVID case reported in that country. The authors showed that no SARS-CoV-2 was detected in samples collected three weeks before the first COVID-19 case; meanwhile, the first virus fragment was detected in sewage at five sites, one week after the first COVID-19 case. More recently, Wu et al. (2020) quantified viral titer of SARS-CoV-2 in sewage from a major urban treatment facility in Massachusetts (USA) and suggested approximately 250 viral particles per mL of sewage. In Queensland (Australia), SARS-CoV-2 RNA tested positive (RT-qPCR) in two out of nine sewage samples, and quantitative estimation were 3–4 orders of magnitude lower than in Wu’s investigation (Ahmed et al., 2020). A reasonable assumption for this discrepancy is the much higher number of COVID-19 infected people in the former region.

These findings show that, despite the lack of knowledge on the persistence of viable SARS-CoV-2 in sewage (WHO, 2020), estimates of its viral load are being scrutinized carefully by health authorities, sanitation operators and scientific community. Accordingly, there is an urgent need for anticipated risk assessment and sanitation interventions in preventing this route of transmission and consequences for public health, considering a possible confirmation of the virus infectivity hypothesis in such environment (Heller et al., 2020). This is particularly important in less developed countries, where the occupational exposure for workers in WWTPs may warrant additional concern since the protocols of personal and collective protective equipment (PPE and CPE) use is not as stringent as it is in the developed countries. Quite recent publications have encouraged the use of QMRA based on previous studies of relevant respiratory viruses, such as SARS-CoV and MERS-CoV, to assess the likely risks of SARS-CoV-2 associated with sewage exposure (Haas, 2020; Kitajima et al., 2020). The focus of this work is to estimate these health risks by incorporating data from the literature, assuming different COVID-19 pandemic exposure scenarios, and by applying a QMRA in WWTP_s_.

## 2. Materials and Methods

### Rationale

QMRA consists of four basic steps: (i) hazard identification; (ii) exposure assessment; (iii) effect assessment (dose-response relationship); and (iv) risk characterization. In occupational settings, these stages should take into account the worker’s activity and identify the transmission chain, routes of exposure, and matrices involved (Carducci et al., 2016).

### Site description

We performed the QMRA with information from two WWTPs from Porto Alegre (South Brazil) - São João Navegantes (SJN-WWTP) and Serraria (S-WWTP). The sewage treatment process at the WWTPs starts with manual (coarse) and automatic (fine) screening, followed by the grit removal. Then, the biological treatment includes an anaerobic stage by the UASB reactor and an aerobic stage through the Unitank^®^ System. The sludge generated in the Unitank^®^ system returns to the UASB reactor and is dehydrated through centrifuges. The effluent is then disposed in the Guaíba Lake, about 1.6 km away from the shore.

We performed the QMRA with information from two WWTPs from Porto Alegre (South Brazil) – São João Navegantes (SJN-WWTP) and Serraria (S-WWTP). The sewage treatment process at the S-WWTPs starts with manual (coarse) and automatic (fine) screening, followed by the grit removal. Then, the biological treatment includes an anaerobic stage by the UASB reactor and an aerobic stage through the Unitank^®^ System. The sludge generated in the Unitank^®^ system returns to the UASB reactor and is dehydrated through centrifuges. The treatment process at the SJS-WWTP includes the same preliminary and final stages, but the biological treatment stage is performed by activated sludge and secondary settling instead of the UASB and Unitank^®^ equipment. The effluent of both WWTPs is then disposed in the Guaíba Lake.

The WWTPs workers perform routine activities of manual cleaning coarse screening, sewage sampling, chemical analyzes, plant inspection and sludge dehydration supervision. In normal bases, WWTP workers wear goggles and gloves as personnel protective equipment (PPE) and treatment units are not cover or equipped with collective protective equipment (CPP) as splashes barriers.

### Hazard identification and exposure assessment

SARS-CoV-2 was considered and chosen for this risk assessment for its primary route to sewage from faeces (Heller et al., 2020), especially in cities that are affected by COVID-19, and potential human ingestion infecting both the intestinal and respiratory tracts. *E. coli*, as one of the most common indicator microorganisms, was also assessed for comparative purposes. The objective of the risk assessment model was to estimate the dose of SARS-CoV-2 to which workers of WWTPs are exposed while performing their work activities. Concentration values of viable SARS-CoV-2 in sewage are not available. In the present study, the concentration of viable (VC) SARS-CoV-2 per mL of sewage (VS) was calculated using Equation 1. This method allocates a diarrhea stool weight (DSW) for western diet of 300 g.d^−1^ (Chandrasekhar, 2020), then multiplies this allocation by the virus concentration in stool of COVID-19 patients (VS) and by the fraction of the population with COVID-19 having diarrhea symptoms (FP). The product is then divided by the flow rate of the WWTP (WF). We assumed the average viral load reported by Lescure et al. (2020) of 6.61×10^7^ genomic copies (GC) of SARS-CoV-2 per g of stool and that 10^3^ genomic copy corresponds to one (1) plaque forming unit - PFU (Aslan et al. 2011;Mcbride et al., 2013; Carducci et al., 2016).

Equation 1

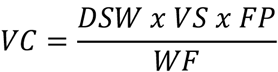

The exposure scenario (Figure 1) was based on the proposed framework for faecal-oral hypothesis raised by Heller et al. (2020) and considered the event of accidental ingestion of sewage by WWTPs workers while performing routine activities. The hazardous exposures were identified by a systematic on-site survey of SJN-WWTP and S-WWTP and their treatment processes and operations. During manual cleaning of coarse screening, when a fork is used to remove wastes, exposure of workers to ingestion of droplets was recognized as the major hazardous event, especially in windy weather conditions, when there is intense contact of sewage droplets and airborne particles with their faces if they are not using a proper PPE as face shield and mask. The volume ingested took into consideration data reported by Westrell et al. (2004) of 1 mL for worker at pre-aeration process. The frequency of exposure was considered to be a single event, since the focus of this study is to assess the risks during COVID-19 outbreaks, rather than extrapolating annual risks.

**Figure 1.**
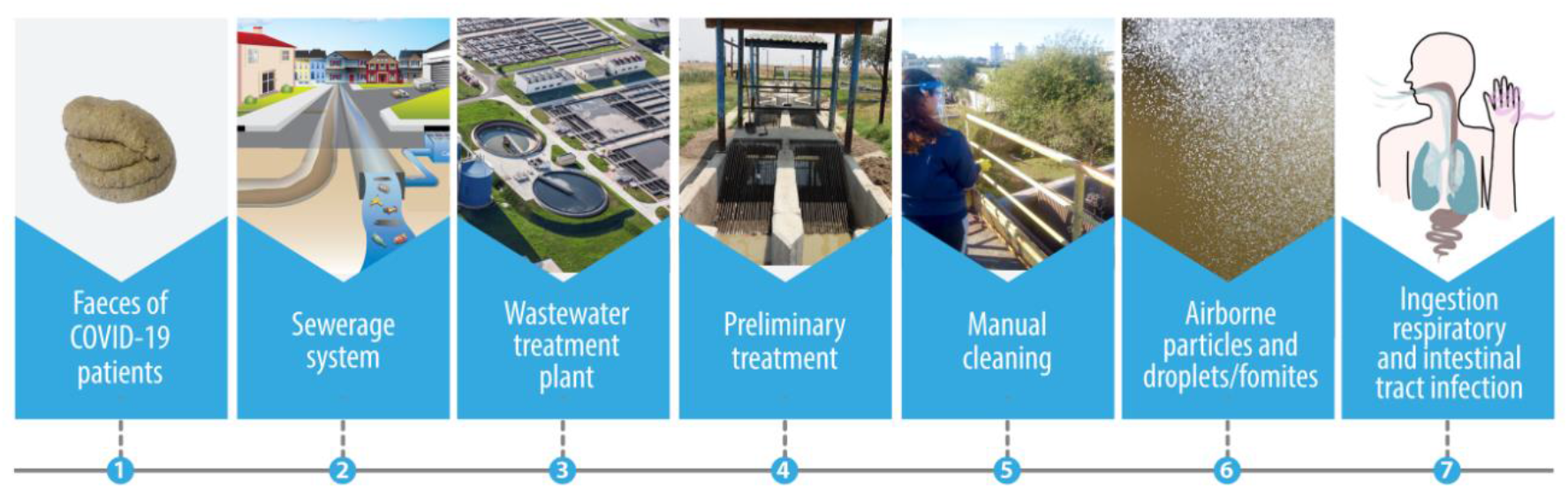
Faecal-oral exposure route of accidental ingestion of sewage by WWTP workers.

A range of three different scenarios were approached, classified according to the total number of COVID-19 infected people, from lowest to highest, as follows: i. moderate, using local data (Porto Alegre, Brazil); ii. aggressive, using data from Madrid (Spain); and iii. extreme, using data from New York City (USA). The evaluation of these three scenarios was divided in 6 types of exposure, three for each WWTP, carried out to study the effect of different stages of the pandemic in terms of percentage of infected population. For estimating total infected population, we used data of COVID-19 infected and recovered people and deaths from April/2020, and accounted for the under-reported cases according to Russell et al. (2020) for New York City and Madrid, and according to UFPEL (2020) for Porto Alegre. The specific values used and sources are described in Table 1.

**Table 1.** QMRA calculations

| Type of exposure | COVID-19<br>infected<br>population in<br>the WWTP<br>contribution<br>area | COVID-19<br>infected<br>population with<br>diarrhea<sup>f</sup> in the<br>WWTP<br>contribution<br>area | SARS-CoV-2<br>viral load in<br>sewage,<br>GC.mL<sup>-1</sup> | Dose (d),<br>PFU<sup>g</sup> or<br>CFU<sup>h</sup> | Risk of<br>infection<br>(P) | Tolerated<br>risk (WHO<br>benchmark<br>value) | Risk of<br>infection (P) /<br>Tolerated risk<br>(WHO<br>benchmark<br>value) |
| --- | --- | --- | --- | --- | --- | --- | --- |
| 1 - SARS-CoV-2 / SJN-WWTP<sup>a</sup> / Extreme<sup>c</sup> | 36,977 | 14,791 | 1.11x10<sup>4</sup> | 11.09 | 2.7E-02 |  | 27 |
| 2 - SARS-CoV-2 / S-WWTP<sup>b</sup> / Extreme<sup>c</sup> | 187,522 | 75,009 | 1.31x10<sup>4</sup> | 13.06 | 3.1E-02 |  | 31 |
| 3 - SARS-CoV-2 / SJN-WWTP<sup>a</sup> / Aggressive<sup>d</sup> | 7,615 | 3,046 | 2.28x10<sup>3</sup> | 2.28 | 5.6E-03 |  | 5.6 |
| 4 - SARS-CoV-2 / S-WWTP<sup>b</sup> / Aggressive<sup>d</sup> | 38,620 | 15,448 | 2.69x10<sup>3</sup> | 2.69 | 6.5E-03 | 1.0E-03 | 6.5 |
| 5 - SARS-CoV-2 / SJN-WWTP<sup>a</sup> / Moderate<sup>e</sup> | 345 | 138 | 1.03x10<sup>2</sup> | 0.10 | 2.5E-04 |  | 0.25 |
| 6 - SARS-CoV-2 / S-WWTP<sup>b</sup> / Moderate<sup>e</sup> | 1,748 | 699 | 1.22x10<sup>2</sup> | 0.12 | 3.0E-04 |  | 0.3 |
| 7 - *E. coli* / SJN-WWTP<sup>a</sup> and S-WWTP<sup>b</sup> | - | - | - | 1x10<sup>4</sup> | 1.06E-03 |  | 1.1 |

<sup>a</sup>Sewage flow rate of 306 L.s<sup>-1</sup>, corresponding to 11.22% of the total population in Porto Alegre (Brazil).
<sup>b</sup>Sewage flow rate of 1,318 L.s<sup>-1</sup>, corresponding to 56.90% of the total population in Porto Alegre (Brazil).
<sup>c</sup>Based on the total estimated population in New York City (United States), in April 2020 of 8,399,000 (United States Census, 2020) and its total COVID-19 active cases of 223,863 (obtained from the difference between the total positive cases of COVID-19 minus the total recovered and deaths (URL 1)). Additionally, not reported COVID-19 cases (88%) were accounted based on Russel (2020), resulting in 22% of the population, in the extreme scenario, infected with COVID-19.
<sup>d</sup>Based on the total estimated population in Madrid (Spain), in April 2020, of 6,642,000 (Eurostat, 2020) and its total COVID-19 active cases of 19,749 (obtained from the difference between the total positive cases of COVID-19 minus the total recovered and deaths (URL 2)). Additionally, not reported COVID-19 cases (93.5%) were accounted based on Russel (2020), resulting in 4.6% of the population in the extreme scenario infected with COVID-19.
<sup>e</sup>Based the total estimated population in Porto Alegre, in April 2020, of 1,483,771 (IBGE, 2020) and its total COVID-19 active cases of 3,072, obtained from UFPEL (2020), considering the under-reported COVID-19 case numbers of 93.2% (UFPEL, 2020), resulting in 0.2% of the total population, in the moderate scenario, infected with COVID-19.
<sup>f</sup>Based on a 40% relation of COVID-19 infected patients with diarrhea symptoms (Lescure et al., 2020).
<sup>g</sup>Unit of dose used for SARS-CoV-2 (type of exposure #1, #2, #3, #4, #5 and #6).
<sup>h</sup>Unit of dose used for *E. coli* (type of exposure #7).

### Dose-response assessment and risk characterization

Because there is no existing dose response model for SARS-CoV-2, a dose-response model for SARS-CoV-1, as a surrogate pathogen, was applied due to the epidemiologic similarities of both coronaviruses in different environments (Doremalen et al., 2020) and the assumptions made by Haas (2020). Watanabe et al. (2010) proposed the exponential model with k = 4.1 × 10^2^ as a dose-response model for SARS-CoV-1 based on the available data sets for infection of transgenic mice susceptible to SARS-CoV-1 and infection of mice with murine hepatitis virus strain 1, which may be a clinically relevant model of SARS. This dose response model was applied in the present work and the risk of infection, in the form of an exponential model, and dose were calculated as in Equations 2 and 3, respectively.

Equation 2

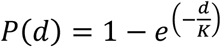

Equation 3

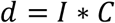

Where:

P=Probability of infection after a single exposure at the dose *d*;
d=Dose, as number of organisms ingested (PFU);
k=dose-response model (4.1 × 10^2^);
I=Volume ingested (1 mL);
C=Concentration of virus in sewage (PFU.mL^−1^).

The dose-response model (Beta-Poisson) and fitting parameters used for *E. coli* risk assessment used herein were the same proposed by Zaneti et al. (2012ab, 2013) for estimating the risk of infection, as in Equation 4. The concentration of *E. coli* at the entrance of the SJN-WWTP and S-WWTP was the average reported from operational data of 10^6^ CFU.100mL^−1^ and we classified this scenario as type of exposure 7# (Table 1), as we assumed that its concentration does not vary in any of the exposure scenarios (moderate, aggressive and extreme). The same exposure frequency (1 single event per year) and volume of sewage ingested (1 mL) used for SARS-CoV-2 were adopted for *E. coli*.

Equation 4

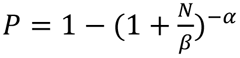

Where:

α=0.1705;
β=1.61×10^6^;
N = Exposure, as number of organisms ingested (CFU).

## 3. Results and discussion

The calculated SARS-CoV-2 viral loads in the raw sewage of the SJN-WWTP and S-WWTP are provided in Table 1. As expected, higher viral loads were observed for the aggressive and extreme scenarios. In the considered SJN-WWTP, the SARS-CoV-2 concentration in sewage was 2.28×10^3^ and 1.11×10^4^ GC.mL^−1^, for aggressive and extreme scenarios, respectively. In the S-WWTP, these values increased to 2.69×10^3^ and 1.31×10^4^ GC.mL^−1^, respectively. This variation in values of about 15% among the WWTPs is attributed to the different flow rates of the WWTPs and the different portions of population that contribute to these plants. In the moderate (local) scenario, the total population infected by COVID-19 were at least 22 times lower than in the other conditions, and viral loads were of 1.03×10^2^ and 1.22×10^2^ GC.mL^−1^ in SJN-WWTP and S-WWTP, respectively.

The assumptions defined in the present work were considered appropriately conservative and protective to human health, since we considered that there is no loss of viral RNA in sewer lines and that excreted viruses are fully suspended in sewage. Despite the uncertainties of such assumptions, the results obtained herein for the moderate scenario are in the same range of the viral titers experimentally determined by Wu et al (2020) in the raw sewage from Massachusetts (USA), which appears to be the first study that directly quantified SARS-CoV-2 in such environment to date. The results obtained for the aggressive and extreme scenario were 1–2 orders of magnitude higher than in Wu’s work, which corroborates with the conservative assumptions made in the present study. The obtained values for the dose ranged from 0.1 to 13.06 PFU, considering the ingested volume of 1 mL. Determining exposure volumes for occupational risks in workplaces such as WWTPs is particularly challenging due to differences in skills and level of experience of workers, the wide range of activities and different levels of protection, as well as seasonal changes. Moreover, there is very little information about the ingestion route during such scenarios. It is generally recognized that accidental ingestion involves the processes of transfer of the substance from the environment into the mouth, and this must include movement of contaminated hands or objects into the mouth, or contact of contaminated hands or objects with the skin around the mouth (the peri-oral area) followed by migration of this contamination into the mouth (Christopher et al. 2006). Splashing into the mouth or onto the face are also relevant mechanisms, although probably much less important. Haas (2020) highlighted the relative importance of the so-called “fomites” in the context of COVID-19 pandemic, which consist of larger airborne particles that can deposit on surfaces, where the contained viruses persist for hours to days, and might have a pathway from hands to mouth, nose and eye. Ashbolt et al. (2005) supported that the volume ingested during operational irrigation activities (water intense activity) follow a triangular distribution of (0.1; 1; 2), in mL. Based on that assumption, we have already carried out another risk-based study on accidental reclaimed wastewater ingestion by using the minimum value (0.1 mL) of this triangular distribution in order not to overestimate the risks (Zaneti et al., 2013; Zaneti et al. 2012ab). On the other hand, the ingestion volume adopted for the proposed exposure pathway in the present study (1 mL in a single event) arose from Westrell et al. (2004), for workers at a pre-aeration treatment stage of WWTP, corresponding to the same value of the peak of the aforementioned triangular distribution of Ashbolt et al. (2005), and thus, not over conservative. Therefore, this approach appears to satisfactorily support the viral loads and doses obtained herein. For the extreme, aggressive and moderate scenarios, the estimated risks reached values up to 3.1×10^−2^, 6.5×10^−3^ and 3.0×10^−4^, respectively. Comparing with the risk assessed for

*E.Coli*, (1.06×10^−3^), considering the same exposure route and extreme scenario, the risks of SARS-CoV-2 are 30 times higher – reinforcing that additional precaution is required during COVID-19 outbreaks. According to Carducci et al. (2018), there are no occupational exposure limits (OELs) for microbial agents to date and thus, acceptable level of risks has not yet been defined. This can make the decision-making process for managing health risks of SARS-CoV-2 in workplaces such as WWTPs a little more complex. In the risk assessment and risk characterization of drinking water and reclaimed wastewater, risk targets are commonly determined in order to set microbial and toxicological limits and develop mitigation strategies (Dogan et al., 2020). Since a focus of this work is to provide a comparison of relative risks, we compared our data to the WHO benchmark of tolerable risk for rotavirus infection in drinking water of 10^−3^ (Mara, 2008) and only in the moderate scenario this benchmark value was not exceeded (Table 1).

WWTP workers are potentially exposed to a variety of infectious agents and toxic materials (Masclaux et al., 2014), but studies on sewage workers exposed to bioaerosols and airborne particles are few. Kitajima et al (2020) reviewed epidemiological key articles on health effects for sewage workers and indicated that three out of four studies noted respiratory and gastrointestinal health impacts as an occupational risk. The present study demonstrated that COVID-19 outbreaks may pose increasing health risks at such workplaces and thus, specific risk management strategies need to be developed. It is highly recommended that wastewater treatment workers that perform manual cleaning of screening use face shields and face masks. At the same time, considering that in most of the emerging countries treatment tanks are not covered in WWTPs, as well as there is no barriers to avoid splashes and sprays as in the developed countries (KDHEKS, 2020; CDC, 2020), it is suggested reduction in circulation (frequency and duration) of workers in such areas. Meanwhile, there are research needs to evaluate experimentally bioaerosol and airborne particle risks for WWTP workers and nearby communities. To date, there is no such work available, but research evaluating SARS-CoV-2 concentration at specific points of the sewer catchment, including the WWTP, as a wastewater-based epidemiology (WBE) strategy seeking to help public health authorities planning epidemic containment is underway (Wu et al., 2020). Such a practice would warn public health officials of community COVID-19 infections earlier than traditional health screenings or virus testing following the onset of symptoms severe enough to warrant medical attention (Hart and Halden, 2020). This early detection could inform an infection risk reduction strategy to mitigate an outbreak.

Our understanding on the potential role of sewage in SARS-CoV-2 transmission is limited by knowledge gaps in its viability in such environment. Nevertheless, the present findings are important to assist stakeholders and WWTPs managers anticipate and reduce an imminent risk and to develop risk management strategies for health protection of workers. In the two evaluated WWTPs of the present study (S-WWTP and SJN-WWTP), security protocols have been strengthened - the use of face shields and masks are now mandatory, and some treatment tanks were covered and/or received barriers to avoid sewage splashes. Looking towards the future, studies combining SARS-CoV-2 molecular detection in sewage and viral isolation in cell culture to confirm infectivity, as well as epidemiological studies are needed to validate these assumptions. Another urgent research need is the risk assessment to communities bordering WWTPs or who simply do not have sewage collection - common situation in underdeveloped countries, which may be subject to routes of exposure to the virus by direct ingestion and inhalation by bioaerosols.

## 4. Conclusions

- An extensive literature review showed that the presence of viral RNA SARS-CoV-2 in stools and sewage has been recently reported and raised concern on a possible faecal-oral transmission of the virus. The incorporation of literature data of SARS-CoV-2, the framework of different exposure scenarios and a dose-response model of a surrogate coronavirus allowed the application of a conservative QMRA of SARS-CoV-2 for workers in WWTPs;
- The estimated viral loads of SARS-CoV-2 in sewage at the entrance of the WWTPs ranged from 1.03×10^2^ to 1.31×10^4^ GC.mL^−1^ (0.1 to 13.06 PFU.mL^−1^, respectively). The QMRA, performed with the aid of a three-tiered approach, showed that only for the moderate scenario the estimated risk of infection for workers was not greater than the WHO benchmark value of 10^−3^. The estimated risk of infection for WWTP_s_ workers were up to 6.5×10^−3^ and 3.1×10^−2^, in the aggressive and extreme scenarios, respectively. These values are higher than that calculated for the *E. coli* risk of infection, demonstrating that conservative risk management strategies, specially the rational use of PPE and CPE, are highly advisable for workers in WWTP^s^.

## Data Availability

All data are available, as referred in the manuscript.

## Acknowledgements

The authors wish to thank DMAE (Municipal Water and Sewage Department from Porto Alegre/RS, Brazil), which allowed the use of operational data and information from WWTPs.

